# Estimating the COVID-19 Prevalence in Spain with Indirect Reporting via Open Surveys

**DOI:** 10.1101/2021.01.29.20248125

**Authors:** Augusto Garcia-Agundez, Oluwasegun Ojo, Harold Hernandez, Carlos Baquero, Davide Frey, Chryssis Georgiou, Mathieu Goessens, Rosa Lillo, Raquel Menezes, Nicolas Nicolaou, Antonio Ortega, Efstathios Stavrakis, Antonio Fernandez Anta

## Abstract

During the initial phases of the COVID-19 pandemic, accurate tracking has proven unfeasible. Initial estimation methods pointed towards case numbers that were much higher than officially reported. In the CoronaSurveys project, we have been addressing this issue using open online surveys with indirect reporting. We compare our estimates with the results of a serology study for Spain, obtaining high correlations (R squared 0.89). In our view, these results strongly support the idea of using open surveys with indirect reporting as a method to broadly sense the progress of a pandemic.

## 1 INTRODUCTION

During the initial phases of the COVID-19 pandemic, progress tracking via massive serology testing has proven to be unfeasible. However, initial estimation methods suggested that the real numbers of COVID-19 cases were significantly higher than those officially reported (1). For instance, by April 30th, 2020, the number of confirmed fatalities due to COVID-19 in the US was 66, 028, and the number of confirmed cases was 1, 080, 303. However, with that number of fatalities the number of cases must have been no less than 4, 784, 637, by simply using the Case-fatality Ratio (CFR) of 1.38% measured in Wuhan (2).

In the case of Spain, the discrepancy seems to be even higher. Preliminary studies point towards only one in 53 cases being reported during the first days of the pandemic (3). Although recent availability of massive testing has reduced this discrepancy, demographic statistics still indicate a degree of underreporting to this day, which can be seen among others in mortality numbers: all-cause mortality statistics in Spain point to two periods of significant excess of deaths in the country over the predicted values in 2020: March and April (44, 599 deaths in excess) and August to December (26, 186 deaths in excess) (4). These numbers contrast with the officially reported number of deaths due to COVID-19, which rests at 50, 837 (5). This discrepancy is corroborated in publications from official government authorities, which indicate an ongoing estimated underreporting of 20% to 40% (6).

In the CoronaSurveys project, (7) we aim to track the progress of the pandemic using online, open, anonymous surveys with indirect reporting. Recent articles have also suggested the use of surveys to monitor the pandemic, both for Spain (8, 9) and globally (10). However, to our knowledge, all surveys conducted in Spain have employed direct reporting only, asking participants about themselves. CoronaSurveys implements the network scale-up method of indirect reporting instead, allowing us to collect data on a wide fraction of the population with a small number of responses and in a very short time-frame (11). In this article, we compare the accuracy of CoronaSurveys with a gold standard: serology testing data collected by the Spanish government in the ENE-COVID study (12).

## 2 METHODS

The survey deployed in the CoronaSurveys project, which can be answered via browser or mobile app, includes two questions:

1. *How many people do you know in your area for which you know their health condition?* The answer to this question by participant *i* is the *Reach r_i_*.
2. *How many of those were diagnosed with or have symptoms of COVID-19?* The answer to this question by participant *i* is the *Cumulative Number of Cases c_i_*.

In the CoronaSurveys project we have focused on simplicity and brevity to maximize interest and retain users that would consistently provide data every few days. For that reason the total number of questions in the survey has been kept small at all times. Our approach yielded good initial results with about 200 responses per week. The survey has been promoted via social networks via direct contacts and, more recently, with paid advertising. To ensure total anonymity, the surveys are hosted on a private instance of LimeSurvey (13). Data is aggregated daily, and in this process the responses are shuffled so no single entry can be back-traced to its user. All the data is published in a public Github repository. The study design was reviewed and approved by the ethics committee of the IMDEA Networks Institute. The survey includes an informed consent.

Once the data is collected, we remove outlier responses. A response is considered an outlier if (1) *r*_*i*_ is outside 1.5 times the interquartile range above the upper quartile (which for the data in this paper means *r*_*i*_ > 175) or if (2) *c*_*i*_/*r*_*i*_ is greater than 1*/*3 (to exclude participants with an exceptionally high contact with cases). For this paper we only consider responses in which participants provide information for their region. Hence, the data is aggregated by region for all participants, to obtain the estimator of COVID-19 prevalence (∑_*i*_ *c*_*i*_)/(∑_*i*_*r*_*i*_) (11).

## 3 RESULTS

To evaluate the accuracy of this method to sense the cumulative number of cases of COVID-19, we compare our estimates with the results of the serology study of Pollán et al. (12) for Spain. We exclude Ceuta and Melilla due to lack of data on our part. Conducted between April 27 and May 11, 2020, the serology study provides data for *n* = 61, 075 participants (0.1787% ± 0.0984% of the regional population, and 0.1299% of the national population). We consider as positive cases those that tested positive to the point-of-care or immunoassay IgG tests (Supplementary Table 6 in Pollán et al. (12), column *Either test positive*).

For our estimates, we consider the (up to) 100 most recent survey responses per region on April 20. The date is chosen because the mean period between illness onset and a 95% confidence of IgG antibodies presence is 14 days (14). This results in *n* = 999 responses (59 ± 35 per region) across Spanish regions, with a cumulative reach of ∑_*i*_ *r*_*i*_ = 67, 199 (0.1827% ± 0.0701% of the regional population, and 0.1434% of the national population).

## 4 DISCUSSION

The Bland-Altman plot in Figure 1A shows a high correlation between the CoronaSurveys estimates and the gold standard. A direct comparison of crude percentages, depicted in Figure 1B, also yields excellent results (*R*^2^ = 0.8994). The linear regression equation points to CoronaSurveys very consistently underestimating the number of cases by a factor of approximately 46%, possibly due to asymptomatic cases. This ratio is consistent with the estimates of the Covid19Impact study of Oliver et al. (9), which used more than 140, 000 direct survey responses collected on March 28th-30th. It is also consistent with the reported data on asymptomatic cases reported by Poll án et al. (12), which found that around a third of the seropositive participants were asymptomatic. Table 1 presents a detailed comparison of the estimates per region obtained in the different studies.

**Table 1.**
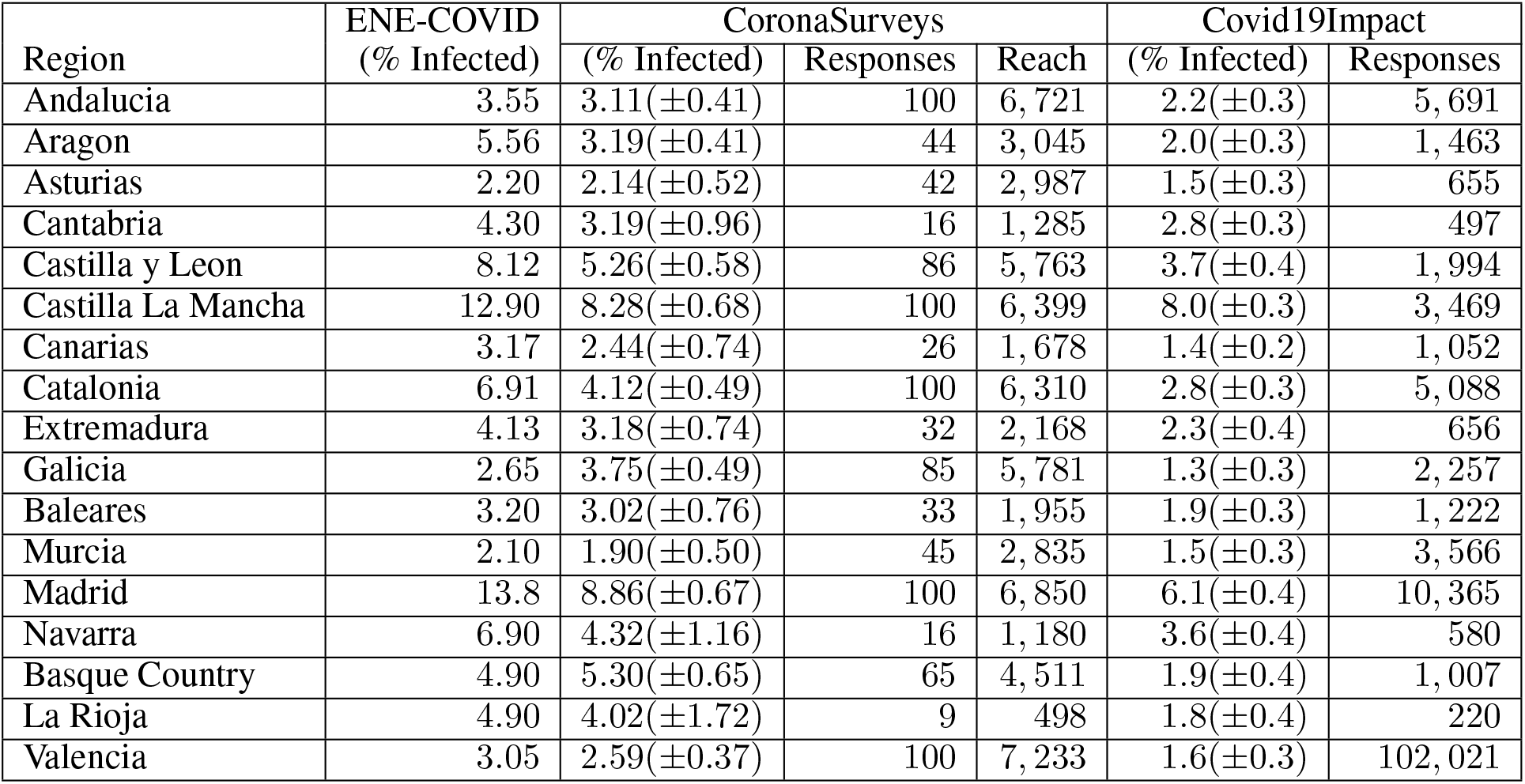
Percentage (and 95% confidence interval) of infected population per region according to the ENE-COVID serology study (12), CoronaSurveys and Covid19Impact (9) (symptom-only model).

**Figure 1.**
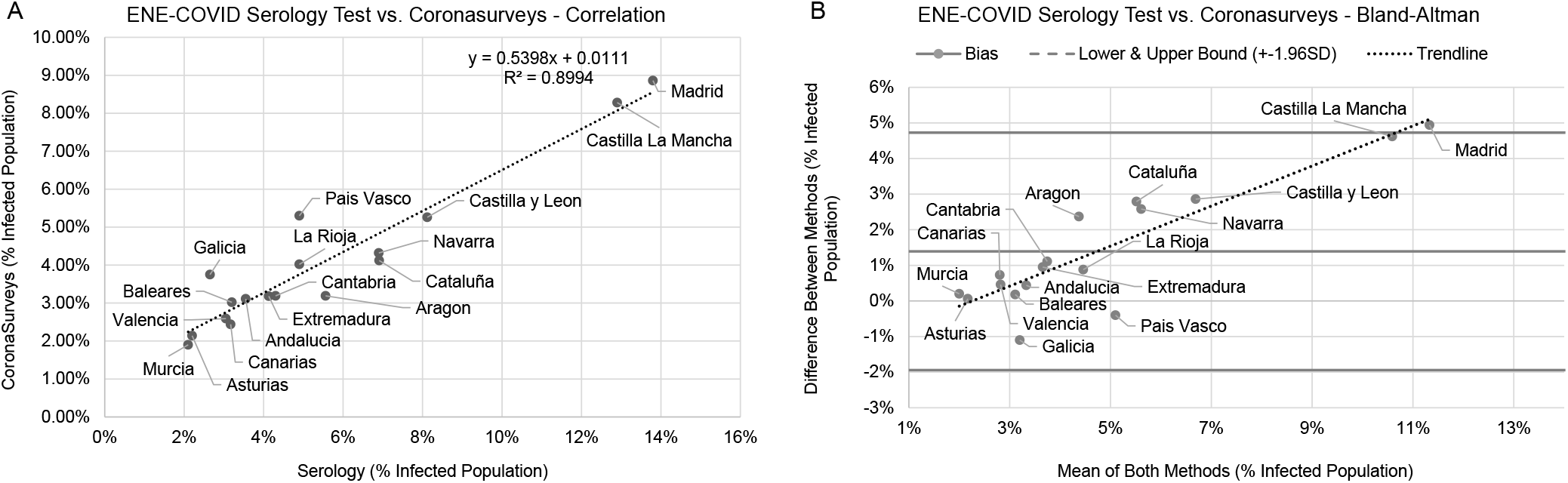
Comparison between the serology test and CoronaSurveys, Bland-Altman (**A**) and direct correlation (**B**)

Figure 2A presents how the number of replies per region affects the resulting value of *R*^2^. This analysis indicates that 50 responses per region can already offer a reasonable estimation of cases. Including more replies may increase accuracy further, but the numbers remain reasonably stable. Naturally, it is important that replies are well distributed across all regions. Figure 2B depicts the effect of the day limit on *R*^2^ if we consider a date of ± one week. Theoretically, a bell curve centered on the 20th should be expected, as estimating too early would imply too few cases are reported, and estimating too late would include more cases. We indeed observe an impact on accuracy, and the left half of the bell curve is more visible. The change in accuracy is mostly due to responses collected on April 16th. The lack of the right half of the bell curve is due to the low number of daily responses after April 16th, which implies that the daily estimates are computed with sets of responses with large intersections.

**Figure 2.**
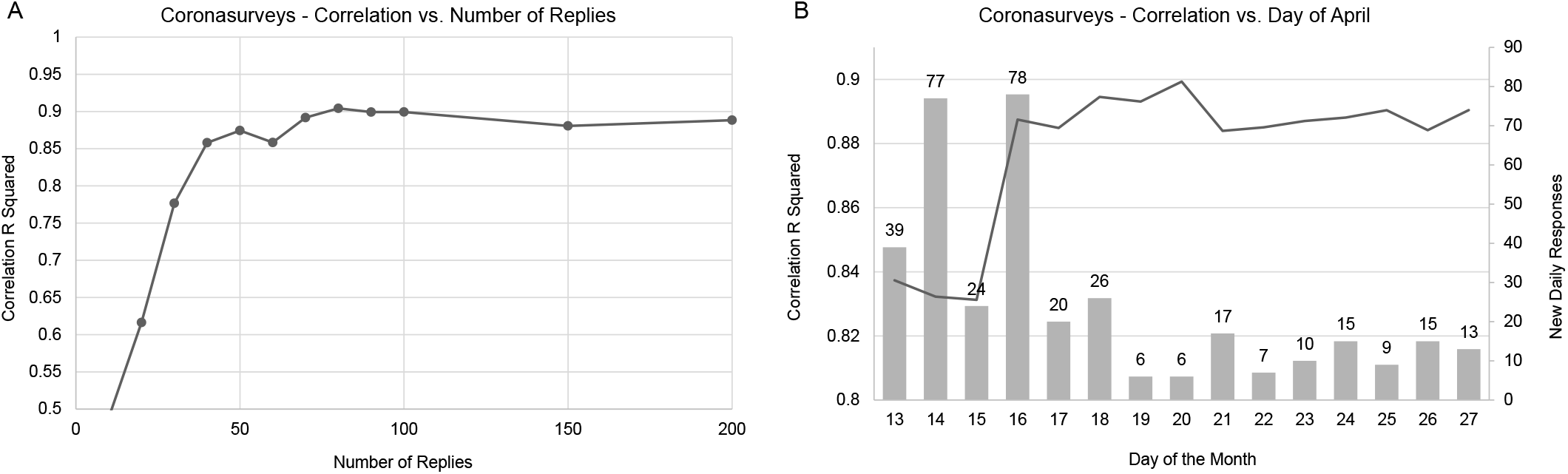
Convergence of correlation with number of replies (**A**) and day of the month (**B**)

Interestingly, a similarly high number of responses was collected on April 14th, with nearly no impact on accuracy. We believe this is due to the distribution of the responses. As depicted in Figure 3, additional responses from regions where many are already available will barely have an impact on the global result. As the great majority of contributions for April 14th were for Madrid, where we already had many responses available, the 77 new responses on April 14th barely had any impact.

**Figure 3.**
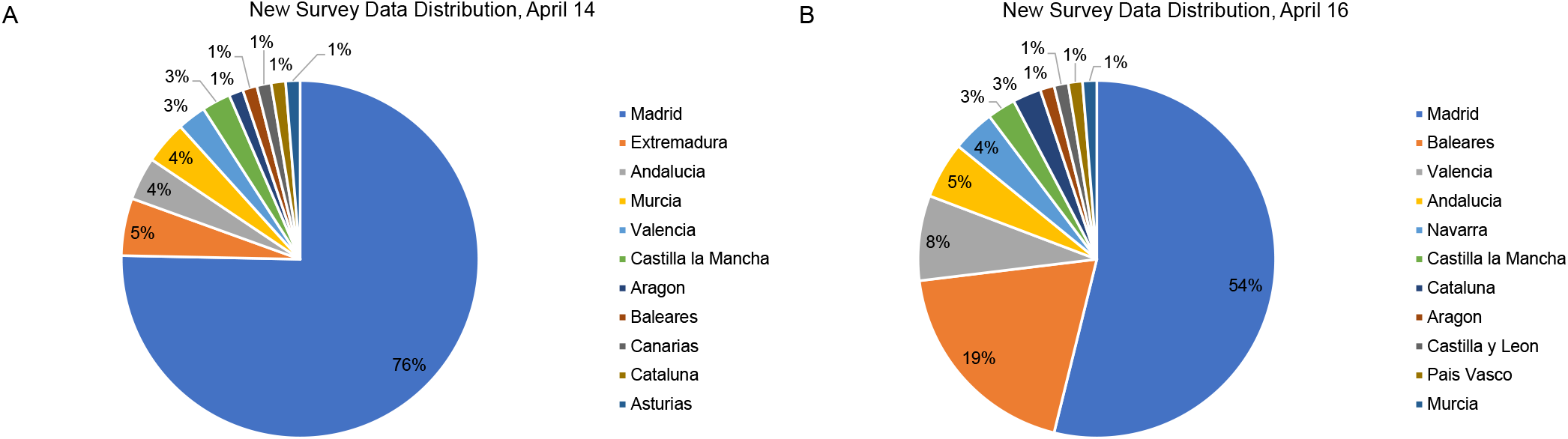
Distribution of new survey responses on April 14 (**A**) and April 16 (**B**)

Our study presents a number of limitations. Firstly, as presented in Table 1, our number of responses in some regions was limited (e.g., 9 responses in La Rioja or 16 in Navarra and Cantabria). Our own analysis suggests this is not enough to offer reliable data for these three regions. Additionally, our criteria to eliminate outliers is heuristic, and may change in the future as we collect more data.

Nevertheless, despite these limitations, the estimates obtained in CoronaSurveys show high correlation with serology tests. Moreover, since the underestimation of our estimates over all regions is homogeneous, and consistent with the one third fraction of asymptomatic reported by Pollán et al. (12), these estimates can be “corrected” to provide an accurate cumulative number of cases for each region. We will further evaluate the robustness of our model as Pollán et al. publish the results of their three additional serology studies.

In summary, we believe these results strongly support using open surveys with indirect reporting as a method to broadly sense the progress of a pandemic.

## Data Availability

All data are publicly available in the CoronaSurveys Github Repository

https://github.com/GCGImdea/coronasurveys

## AUTHOR CONTRIBUTIONS

The analysis presented in this article was conducted by Augusto Garcia-Agundez and Antonio Fernandez Anta with support and feedback from all remaining co-authors. The data acquisition and processing techniques were developed by all co-authors.

## ACKNOWLEDGMENTS

We would like to thank all CoronaSurveys researchers and collaborators for their contribution to this project: https://coronasurveys.org/team/.

## DATA AVAILABILITY STATEMENT

The datasets generated and analyzed for this study can be found in the CoronaSurveys Github Repository at https://github.com/GCGImdea/coronasurveys.

